# Modelling the transmission of infectious diseases inside hospital bays: implications for Covid-19

**DOI:** 10.1101/2020.09.04.20188110

**Authors:** David Moreno Martos, Benjamin J. Parcell, Raluca Eftimie

## Abstract

Healthcare associated transmission of viral infections is a major problem that has significant economic costs and can lead to loss of life. Infections with the highly contagious SARS-CoV-2 virus have been shown to have a high prevalence in hospitals around the world. The spread of this virus might be impacted by the density of patients inside hospital bays. To investigate this aspect, in this study we consider a mathematical modelling and computational approach to describe the spread of SARS-CoV-2 among hospitalised patients. We focus on 4-bed bays and 6-bed bays, which are commonly used to accommodate various non-Covid-19 patients in many hospitals across UK. We use this mathematical model to investigate the spread of SARS-CoV-2 infections among patients in non-Covid bays, in the context of various scenarios: changes in the number of contacts with infected patients and staff, having symptomatic vs. asymptomatic patients, removing infected individuals from these hospital bays once they are known to be infected, and the role of periodic testing of hospitalised patients. Our results show that 4-bed bays reduce the spread of SARS-CoV-2 compared to 6-bed bays. Moreover, we show that the position of a new (not infected) patient in specific beds in a 6-bed bay might also slow the spread of the disease. Finally, we propose that regular SARS-CoV-2 testing of hospitalised patients would allow appropriate placement of infected patients in specific (Covid-only) hospital bays.

## 1. Introduction

There can be high mortality and morbidity for patients due to the spread of infections in hospitals (i.e., nosocomial or healthcare-associated infections) from infected patients to healthy patients, either directly through sharing the same rooms and environment, or indirectly through healthcare workers (HCWs) attending infected and susceptible patients. The novel coronavirus disease (Covid-19) that has been reported since January 2020, was shown to cause a relatively high prevalence of nosocomial infections: between 12.5 − 15% in the UK [4, 42] and up to 44% in China [55]. In comparison, other coronaviruses such as SARS (Severe Acute Respiratory Syndrome, 2003) and MERS (Middle East Respiratory Syndrome, 2012), had estimated nosocomial infection prevalence of 36% and 56%, respectively [4].

In UK hospitals, patients symptoms and results of real-time polymerase chain reaction (RT-PCR) inform targeted infection prevention and control (IPC) measures [42, 4]. Patients who do not meet the clinical criteria for the COVID-19 disease may not be tested, and are usually admitted into shared bays, i.e., rooms of up to 6 beds that usually share the same bathroom facilities [42]. There can be problems with this approach, as highlighted in a recent study [42] in which it was shown that more than half of the patients who acquired Covid-19 in a hospital setting were identified as having been in the same hospital bay with other infected patients.

There is also the issue of asymptomatic cases, which pose a particular risk for healthcare transmission. Various studies in the literature have estimated the proportion of asymptomatic Covid-19 infections: from 17.9% asymptomatic passengers on board of the Princess Cruises ship [31], to 30.8% asymptomatic Japanese nationals evacuated from Wuhan [35], 50–75% asymptomatic residents of an Italian village (Vo’Euganeo) [9], and even 81% asymptomatic passengers of an un-named cruise ship [20]. Since asymptomatic infections cannot be recognised if they are not confirmed by laboratory tests [35], it is highly likely that asymptomatic patients are found in hospitals, and they can be involved in the spread of Covid-19 inside these hospitals. In fact, a study in Beijing [49] showed that 5% of hospitalised patients were asymptomatic. This complicates the issue of nosocomial transmission of Covid-19, since hospitals do not test/re-test routinely all patients hospitalised for various interventions (not Covid-related).

In regard to nosocomial transmissions, a 2015 report by the National Institute for Health Research [29] concluded that while hospital patients preferred a single-room accommodation, staff preferred a mix of single-rooms and shared-rooms accommodation. Moreover, a study examining hospital design [29] mentioned that it was impossible to perform an analysis of the impact of single-room hospital design on the transmission of hospital-acquired infections, and given that some of the studies they cited showed a decrease in infections while other studies showed an increase in infections, the authors concluded that there was no clear evidence of the impact of single rooms on hospital acquired infections. Since some of the published Covid-19 studies [42] emphasised that *≈* 55% of hospital-acquired infections were connected with infected patients in shared bays, it is important to understand Covid-19 transmission in these hospital bays.

The common limitation of clinical studies that focus on the spread of Covid-19 inside hospitals (see, for example, [4, 42, 52]), is that they identify and count only PCR-symptomatic patients and not hospitalised patients that were exposed to the disease but were discharged before developing symptoms. Also, these studies cannot identify contacts between patients and staff, which might have had undiagnosed Covid-19 [42]. To address this lack of information, mathematical modelling and computational simulations are the perfect tool to test various scenarios regarding the mechanisms that can reduce the Covid-19 transmission in hospitals. Currently there are very few mathematical studies that focus on the nosocomial transmission of SARS-CoV-2 (despite the huge number of studies focused on the general viral spread through communities; see, for example, [18, 21, 28, 16, 39, 7, 14, 36, 48, 53, 37, 23, 25, 5, 38, 24, 8, 51, 3, 11, 47, 2] and references therein). We are aware of only one modelling study in [13], where the authors used a classical SEIR continuous model to investigate the transmission dynamics of COVID-19 in English hospitals, following interactions between patients and health-care workers.

Here we present a new study that considers a mathematical modelling approach to investigate the spread of Covid-19 among patients hospitalised in 4-bed versus 6-bed bays, which represents a standard bay structure in many hospitals in the UK; see also Figure 1. To this end we consider a network model that accounts for stochastic fluctuations between interconnected individuals inside the same hospital bay, where every single bed can be occupied by a susceptible, exposed, infectious or a recovered individual. With the help of this model we investigate computationally a variety of scenarios: from variations in the recovery and incubation periods; to the position of infected individuals in specific beds inside 6-bed bays; the effect of removing infected individuals from these bays; the effect of accommodating patients in single rooms; the effect of having asymptomatic individuals; and the role of periodic testing of hospitalised patients.

**Figure 1:**
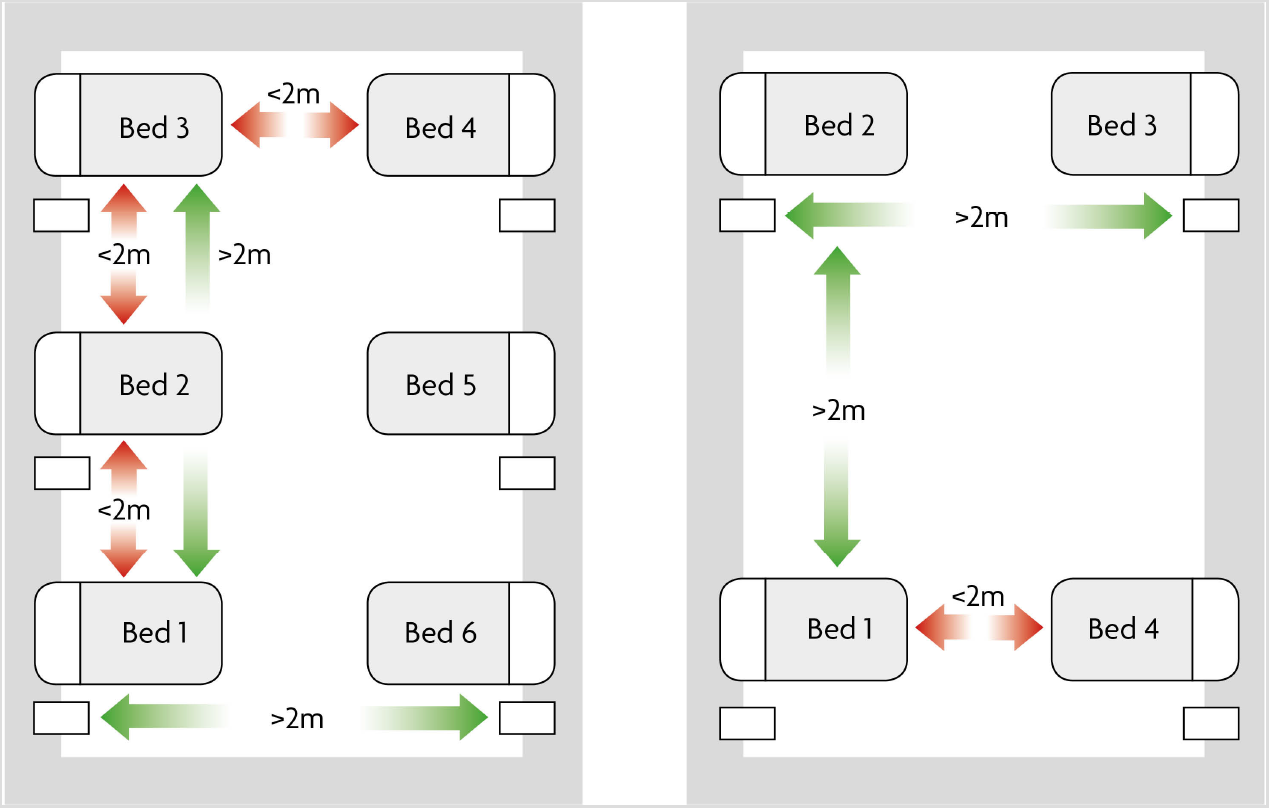
Typical distribution of beds in UK hospitals. Left: 6-bed bays; Right: 4-bed bays. In red it can be seen the distance between beds less than 2m, while in green the distance between beds greater than 2m.

## 2. Materials and method

### 2.1. Model description

Since we are interested in investigating the transmission of infectious diseases (including Covid-19) across 4-bed and 6-bed hospital bays, which are the standard bays in many UK hospitals, in the following we consider a individual-based (network) model that tracks susceptible (S), exposed (E), infected (I) and recovered (R) individuals in each of the 4 or 6 beds inside these hospital bays. Following the approach in [33], each individual *I*_*n,t*_ is described by a set of characteristics, *I*_*n,t*_ = [*C*_*n*,1,*t*_, *C*_*n*,2,*t*_,…, *C*_*n,m,t*_], where *n* = 1*,…N* describes the number of individuals in the hospital bay, *m* is the number of characteristics, and *t* is the time. These characteristics are as follows:

- *C_n,1,t_* is the epidemiological class. It can be 0 (susceptible), 1 (exposed), 2 (infected) or 3 (recovered).
- *C_n,_*_2*,t*_ is the bed at which the individual is placed.
- *C_n,_*_3*,t*_ is the time since the individual acquired the virus (start of the incubation period).
- *C_n,_*_4*,t*_ is the duration of the incubation period.
- *C_n,_*_5*,t*_ is the time since the individual has become infectious (virus transmission is now possible).
- *C_n,_*_6*,t*_ is the time that takes the individual to recover once becomes infectious.

The following rules are used to update each of these states:

- *Epidemiological class update:*

– A susceptible individual might become exposed with probability *β* if he/she interacts with an infected individual. Throughout this study we assume that the infection probability *β* depends on the distance between the susceptible individual and the infectious one. This modelling assumption is consistent with the clinical assumption of predominant transmission of SARS-Cov-2 via respiratory droplets [43]. Some recent studies have suggested that airborne transmission may be possible in certain circumstances, however to date the authors report that there is no perfect experimental data proving or disproving airborne transmission of SARS-CoV-2 [22]. Moreover, the evidence seems inconsistent with airborne transmission especially in well-ventilated spaces [22]. For this reason, and in accordance with current public health recommendations [40], in this study we assume that a distance of at least 2m distance should be maintained to minimise the spread of SARS-CoV-2. Thus, here we consider the infection probability *β* to be a fixed number that depends on the distance between beds: probability is 1 if the distance is 0, probability is almost 0 if the distance is greater than 2; see the exponential distribution in Figure 2. The distance *d_ij_* between two beds *i* and *j* (see also Figure 1) is given by matrices *D_k_* = (*d_ij_*), with *i, j* = 1*..k* and *k ∈ {*4, 6*}*:

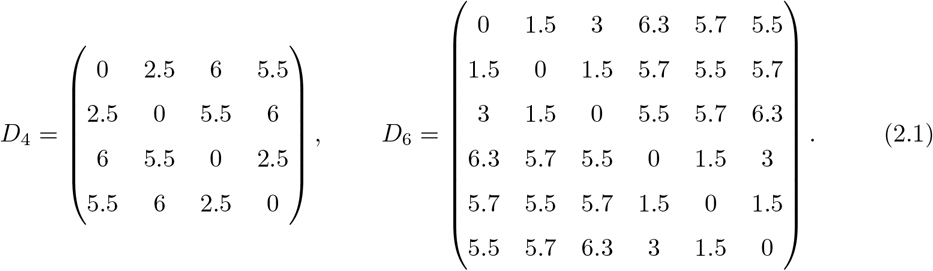 Therefore, the probability of becoming infected following the “contact” between individuals in beds *i* and *j* (where “contact” is understood in a very loose sense) is *β*(*d_ij_*); see alsoFigure 2.
– An exposed individual becomes infected once his/her incubation period is finished (i.e., *C_n,3,t_* > *Cn,_4,t_*).
– An infected individual recovers if the time since he/she has been infected surpasses the expected duration of the infection (i.e., *C_n,_*_5*,t*_ > *C_n,6,t_*).
- *Incubation time update:* If the individual is exposed, the incubation time update is increased by ∆*t* at each iteration, until the individual becomes infectious.
- *Incubation stage:* We describe the duration of the incubation stage using an Erlang distribution with mean 1*/σ* and shape 2. Since the results obtained with this Erlang distribution are relatively similar to the results obtained with an exponential distribution (not shown here; for details see [32]), throughout this study we use only Erlang distributions.
- *Infection time update:* If the individual is infected, the infection time update is increased by ∆*t* at each iteration, until the individual recovers.
- *Recovery stage:* Similar to the incubation stage, to describe the duration of the recovery stage we use an Erlang distribution with mean 1*/γ* and shape 2.

**Figure 2:**
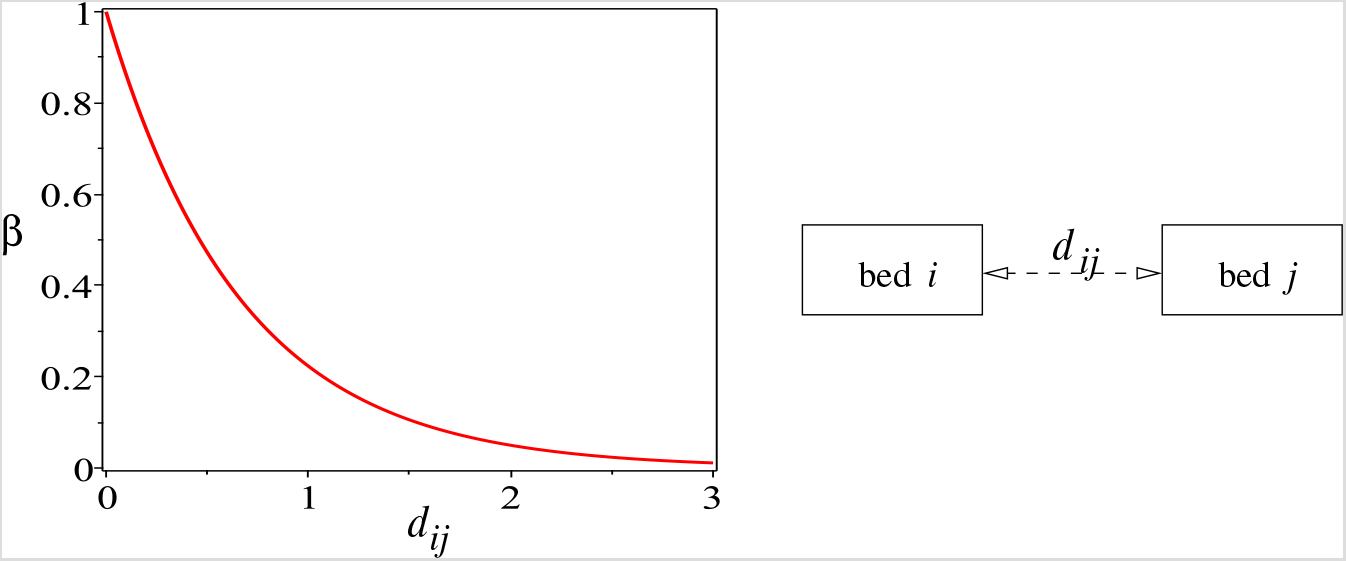
Probability distribution of infection depending on the distance *d_ij_* between beds *i* and *j*: 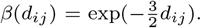. It is assumed here that the infectious disease spreads mainly though respiratory droplets, which fall to the ground rapidly within 1–2m of the source person [22]. The case of airborne transmission, where aerosol particles persist in the air, can be modelled by a similar exponential function with a very low coefficient for *d_ij_* – not investigated in this study, as explained in the main text.

Before we show the numerical results, we needs to discuss briefly the parameter values for which these results are obtained. Even if this model can be generally applied to the spread of various infectious diseases through hospital bays, here we focus on the spread of Covid-19. Therefore, the parameter values (i.e., mean recovery period, mean incubation period) are chosen for this particular disease, Covid-19, and are summarised in Table 1.

**Table 1:** Summary of model parameters used throughout this study. In the 4^*th*^column we show the most common values and ranges for the parameters that were found in the published literature (see references listed in the 5^*th*^column).

| Parameters<br>(& units) | Description | Range | Most common<br>values | References |
| --- | --- | --- | --- | --- |
| $\sigma$ (day<sup>-1</sup>) | Incubation rate | 0.071 – 0.5 | 0.143, 0.192 | [28, 16, 39, 7, 14, 36, 48, 53, 37, 23, 25, 5, 38, 24, 8, 51, 3, 11, 47, 2] |
| $1/\sigma$ (day) | Mean incubation period | 2 – 14 | 7, 5.2 | [28, 16, 39, 7, 14, 36, 48, 53, 37, 23, 25, 5, 38, 24, 8, 51, 3, 11, 47, 2] |
| $\gamma$ (day<sup>-1</sup>) | Recovery rate | 0.055 – 0.621 | 0.143 – 0.25 | [28, 34, 41, 16, 39, 7, 14, 36, 48, 53, 5, 38, 24, 51, 3, 11, 47, 2, 26] |
| $1/\gamma$ (day) | Mean recovery / infection period | 1.61 – 18 | 4 – 7 | [28, 34, 41, 16, 39, 7, 14, 36, 48, 53, 5, 38, 24, 51, 3, 11, 47, 2, 26] |

## 3. Results

For the numerical simulations presented here, we fix 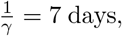 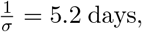 and we run the simulations up to *T_max_* = 60 days. We run this network model 300 times, and we compute the mean and standard deviation of these 300 runs for individuals in each epidemiological state (i.e., susceptibles, exposed, infectives, recovered), for each of the 4-beds or 6-beds bays. We already mentioned before that the probability of infection *β* is given by a decaying exponential which depends on the distance between beds (see Figure 2). Finally, as initial conditions for these simulations, we assume that all patients are susceptible, with the exception of the individual placed in bed 2 (see Figure 1), who is exposed. This describes the situation where an individual, not thought to have the disease (although they were exposed), is introduced into a hospital bay full of patients that can become infected.

### 3.1. Baseline simulation results

We start this numerical investigation of our network model by assuming that hospitalised patients are accommodated in shared bays with either 4 beds or 6 beds. In Figure 3 we show the baseline dynamics of this model when an infected patient is hospitalised in Bed 2 of a: (a),(b) 4-bed bay; (c),(d) 6-bed bay. We observe that in a 4-bed bay the average number of exposed and infected individuals decreases towards zero faster than in a 6-bed bay (see panels (a),(c)). Moreover, we note that for a 4-bed bay, individuals placed in Beds 3 and 4 have an almost zero probability of becoming infected, when the original exposed patient was placed in Bed 2 (see Figure 3(b)). For a 6-bed bay, individuals placed in Beds 4, 5, and 6 have an almost zero probability of becoming infected, when the original exposed patient was placed in Bed 2 (see Figure 3(d)). This means that the distance between the infected Bed 2 and the beds on the opposite walls is large enough to avoid the infection.

**Figure 3:**
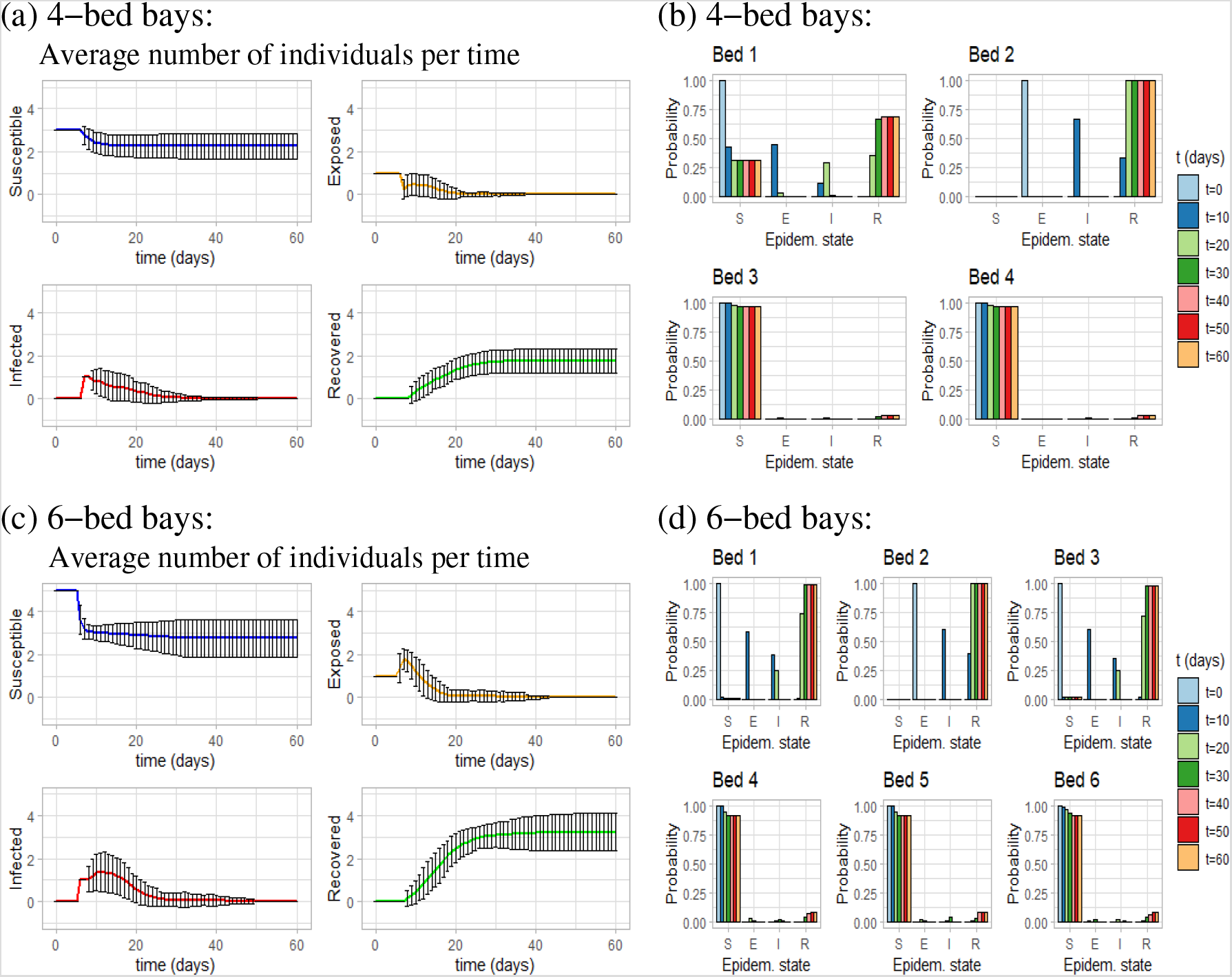
Baseline dynamics for the network epidemiological model introduced in this study. (a) Average number of individuals per time, in a 4-bed bay; (b) Probability of being at each epidemiological state per bed, in a 4-bed bay; (c) Average number of individuals per time, in a 6-bed bay; (d) Probability of being at each epidemiological state per bed, in a 6-bed bay. Here we assume that 1*/γ* = 7 days, 1*/σ* = 5.2 days, ∆*t* = 0.1, and *T_max_* = 60 days.

### 3.2. Placing the initial exposed individual in different beds

In a 4-bed bay it does not matter where we place the first exposed individual, as the disease spreads equally in all beds due to the symmetry of bed distribution. However, in a 6-bed bay, it is important where we place the exposed individual: the disease is more likely to reach all the individuals in a room if the exposed individual is placed in beds 2 or 5. Next we compare how the disease evolves depending on where we place the first exposed individual in a 6-bed bay.

In Figure 4 we assume that the first exposed individual is placed in Bed 1, which is different from the case in Figure 3(b),(d) where we assumed that the first exposed individual was placed in Bed 2. We see that by placing the first exposed individual at Bed 1, the disease spreads a bit slower compared to the case when this individual is placed in Bed 2. More precisely, when we place him/her in Bed 1, the average number of susceptible individuals decreases and reaches a steady value around *t* = 35 days, whereas when we place him/her in Bed 2 the steady value is reached around *t* = 28 days.

**Figure 4:**
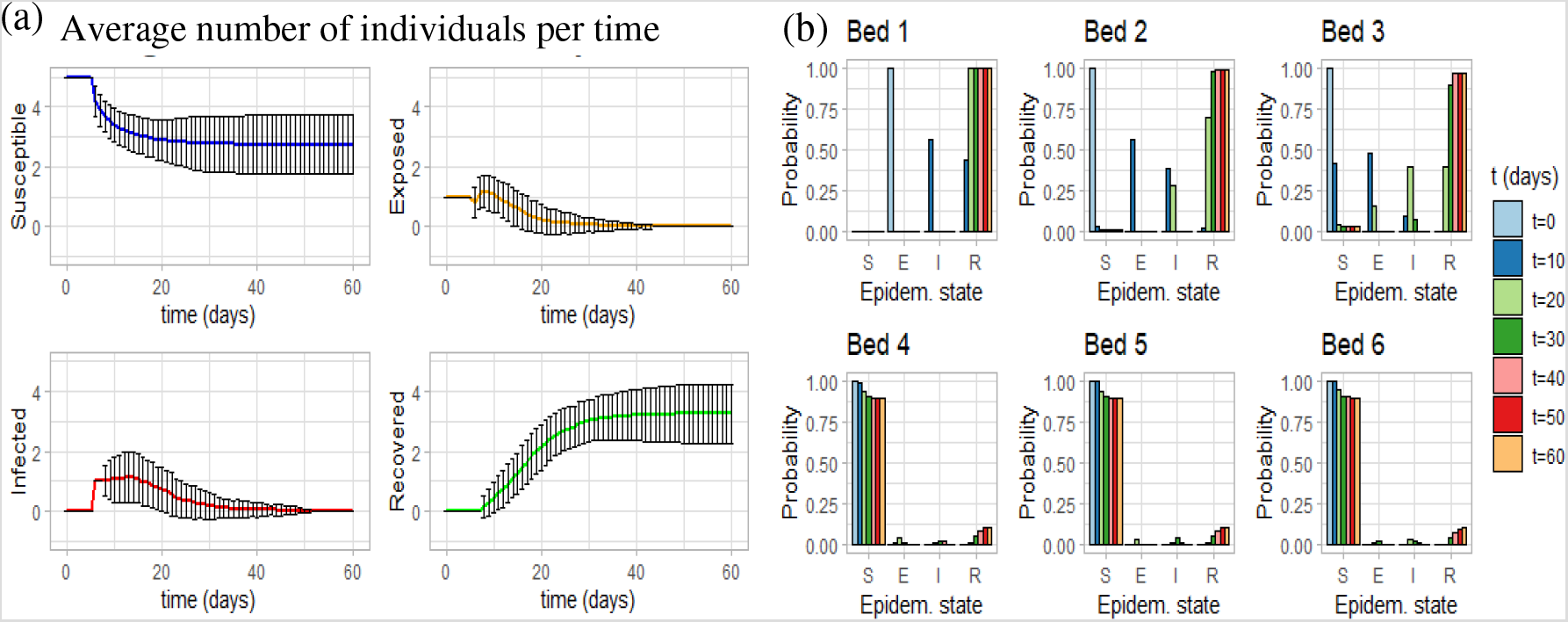
The spread of infection in a 6-bed bay when the initial exposed individual is placed in Bed 1. (a) Average number of individuals per time. (b) Probability of being at each epidemiological state per bed. We assume that 1*/γ* = 7 days, 1*/σ* = 5.2 days, *T_max_* = 60 days, and Δ*t* = 0.1.

### 3.3. Varying the recovery and incubation periods

The parameters that characterise the transmission of Covid-19, e.g., mean incubation period and mean recovery period, are not fixed, with various studies showing different estimations; see Table 1. Here we focus on a 6-bed bay, and investigate the impact of incubation and recovery rates, showing only results for the minimum and the maximum values of these parameters. When varying the incubation rate we fix the mean recovery period to 1*/ γ* = 7 days, and when varying the recovery rate we fix the mean incubation period to 1*/ σ* = 5.2 days. (Here we do not vary *β*, since this parameter is given by the distance between beds; see caption of Figure 2.)

Figure 5 shows model dynamics when we modify the recovery period. The most significant difference between the cases 1*/γ* = 1.61 days and 1*/γ* = 18 days is in the amplitude and shape of the infected curve. For the case where the recovery period is 18 days (Figure 5(c)) there is a time interval during which the number of infected individuals reaches a value of 2 (within one standard deviation). Since the recovery periods are larger (the mean recovery period is 18 days), there is more time in which susceptible individuals can become infected. This also affects the values of the average number of susceptible and recovered individuals. In terms of the probability of being at each epidemiological state per bed, there is a higher probability of becoming infected when the mean recovery period is 18 days (panels (d)), due to larger exposure to infected individuals.

**Figure 5:**
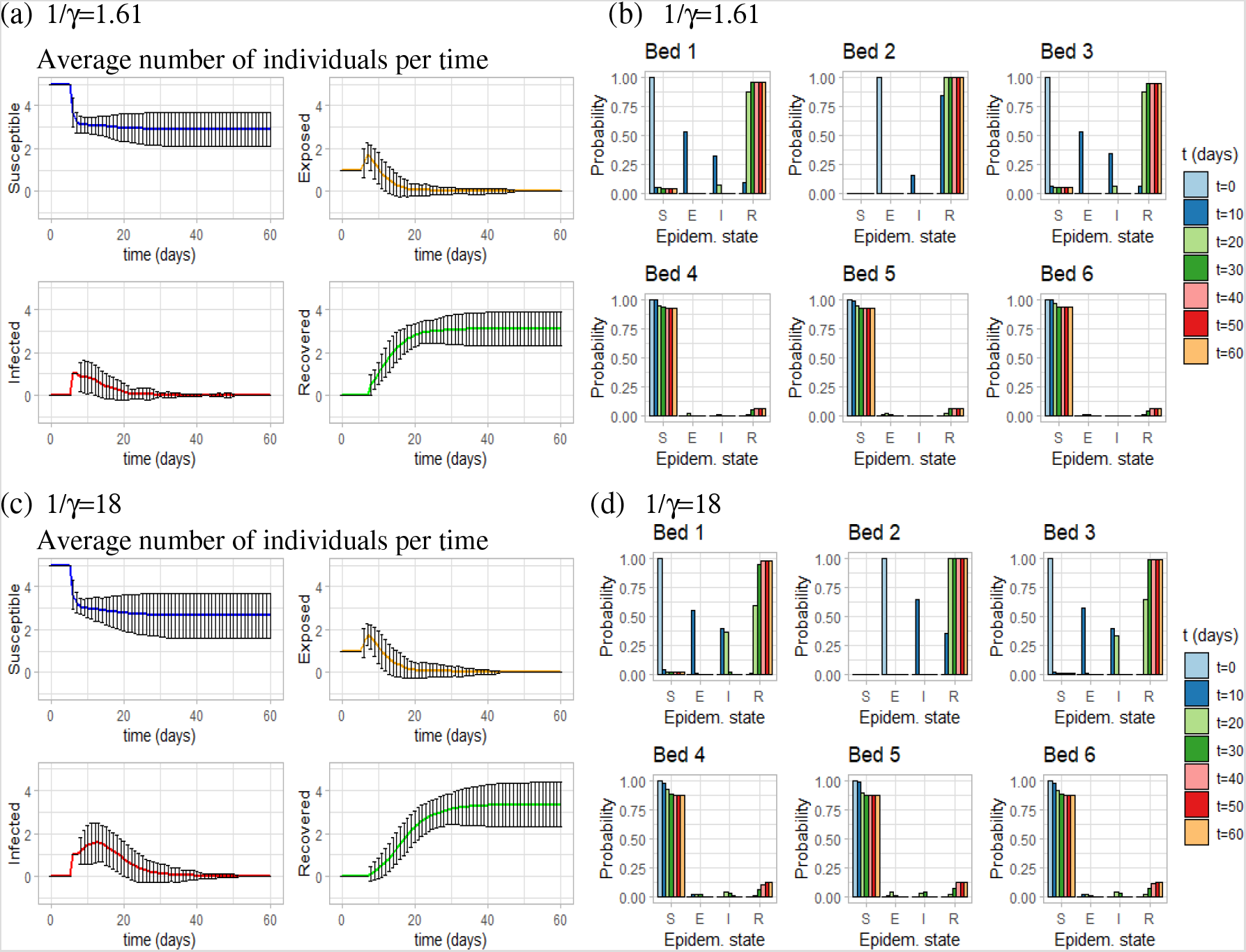
Disease spread across a 6-bed bay, when we fix 1*/σ* = 5.2 days and we vary 1*/γ ∈* (1.61, 18) days. (a) Average number of individuals per time when 1*/γ* = 1.61 days; (b) Probability of being at each epidemiological state per bed, when 1*/γ* = 1.61 days; (c) Average number of individuals per time when 1*/γ* = 18 days; (d) Probability of being at each epidemiological state per bed, when 1*/γ* = 18 days. As before we run simulations for *T_max_* = 60 days, and Δ*t* = 0.1.

The effects of varying the incubation period are shown inFigure 6. The most significant differences between the cases 1*/σ* = 2 days and 1*/σ* = 14 days can be found in the average numbers of exposed and infected individuals. When the mean incubation period is longer (Figure 6(c)), the mean value for the exposed and infected individuals reaches zero later in time. In terms of the probability of being at each epidemiological state per bed, the probability of being infected is higher for earlier times (*t* = 10 days), when the mean incubation period is shorter (panels (c)).

**Figure 6:**
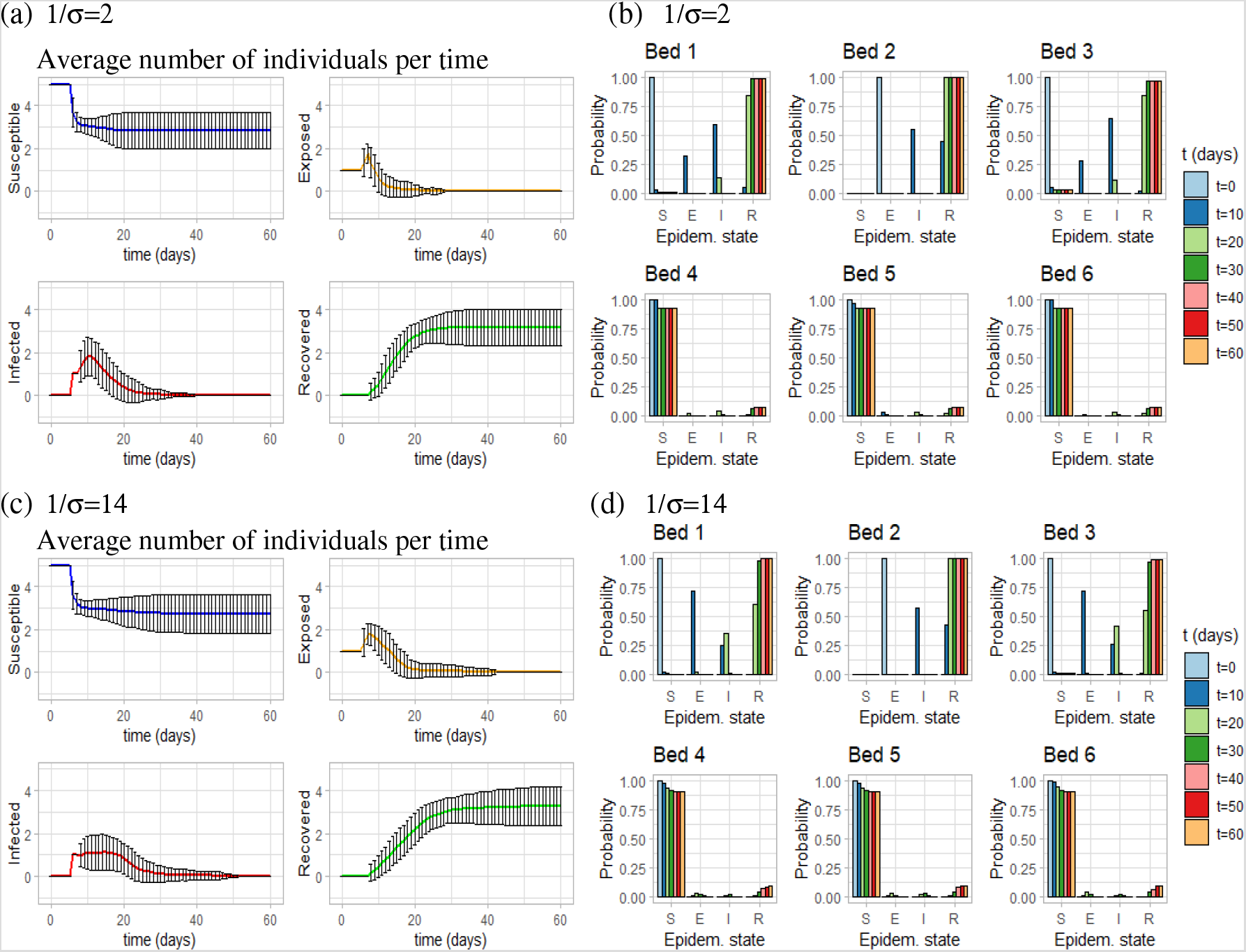
Disease spread across a 6-bed bay, when we fix 1*/γ* = 7 days and we vary 1*/σ ∈* (2, 14) days. (a) Average number of individuals per time when 1*/σ* = 2 days; (b) Probability of being at each epidemiological state per bed, when 1*/σ* = 2 days; (c) Average number of individuals per time when 1*/σ* = 14 days; (d) Probability of being at each epidemiological state per bed, when 1*/σ* = 14 days. As before we run simulations for *T_max_* = 60 days, and Δ*t* = 0.1.

### 3.4. The effect of removing infected individuals from the bays

Here we investigate the effect of removing from the bay every patient that is classified as infected (we remove this patient when the incubation period ends). We do this in order to reproduce the situation in which patients are transferred to separate Covid-19-only bays right at the moment they present clinical symptoms (assuming all of them are symptomatic individuals). Once we remove the infected patient, we place a new susceptible individual in the free bed. We emphasise that we modelled this out of mathematical interest (to keep the same number of patients in a bay and continue the simulations), as the standard clinical practice does not put a new person back in a bay with potential incubating patients.

Since various studies reported pre-symptomatic transmission of SARS-CoV-2 during the incubation period [50, 45, 54], and following the approach in some published modelling studies [28, 37] which accounted for the spread of infection during the latent period, in the following we assume that exposed individuals can transmit the disease, but at a much lower rate compared to the infected ones (we consider here an overlap between exposed and pre-symptomatic cases). Since we could not find in the literature any values for the probability of transmission, in this sub-section we set the probability of infection from the exposed individuals to be 20% of the probability of infection from the infected individuals. In Figure 7 we can see the differences between 4-bed bays and 6-bed bays. On one hand, in a 4-bed bay the disease disappears after a month. This is when we have 4 susceptible and 0 exposed in all the runs (Figure 7(a)) or the probability of being in the exposed state is zero for times greater than 30 days (Figure 7(b)). On the other hand, in a 6-bed bay the disease disappears after 50 days, which is when the probability of being in the exposed state is zero for *t >* 50 days (Figure 7(d)). Both bays have in common the fact that the individuals across the aisle from the infected Bed 2 (i.e., Beds 3 and 4 in a 4-bed bay and Beds 4, 5 and 6 in a 6-bed bay) do not become infected.

**Figure 7:**
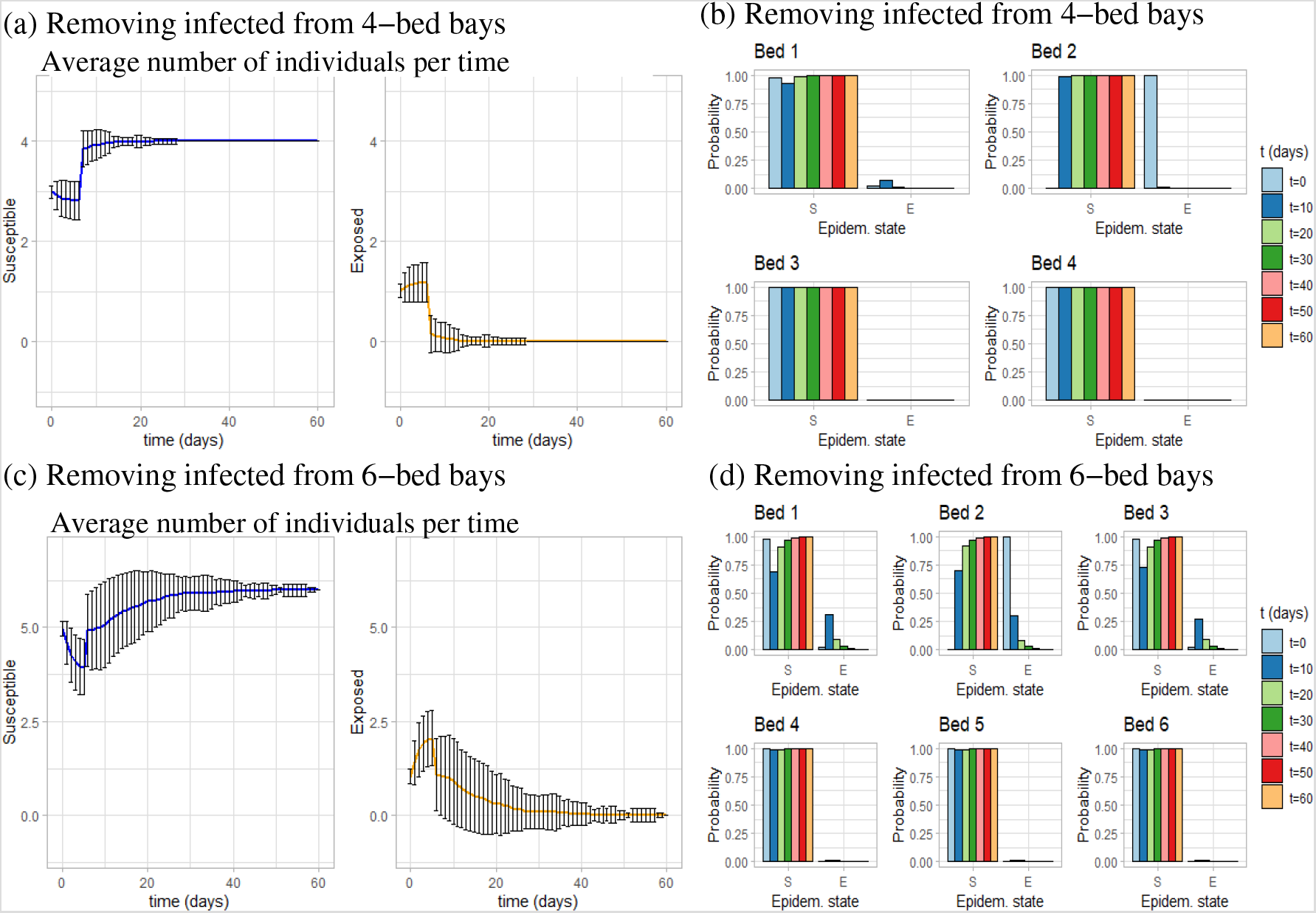
Spread of the disease when infected individuals are removed from (a),(b) 4-bed bays; (c),(d) 6-bed bays. (a) Average number of individuals per time, in a 4-bed bay; (b) Probability of being at each epidemiological state, in a 4-bed bay. (c) Average number of individuals per time, in a 6-bed bay; (b) Probability of being at each epidemiological state, in a 6-bed bay. Here we assume that 1*/γ* = 7 days, 1*/σ* = 5.2 days, *T_max_* = 60 days, and Δ*t* = 0.1.

### 3.5. Effect of interacting with members of the staff and sharing bathrooms

Until now, we have assumed that patients in a hospital bay can become infected by interacting with exposed/infected individuals from the same bay. However, sometimes they have to share bathrooms with individuals from other bays, which increases the probability of becoming infected (it has been shown that the virus can survive for several hours on different surfaces [12]). Moreover, patients interact with hospital staff (doctors, nurses, etc.), who might transmit the disease with a very low probability [46], even if they follow the preventive measures (washing their hands, changing their clothes, wearing masks, etc.).

To implement this aspect in our simulations, we note that the secondary attack rates for Covid-19 (defined as the probability that infection occurs among susceptibles, following contacts with an infectious person or infectious environment [17]) have an estimated range between 0.46%–63.87% [19]. In a study focused on Taiwan patients [19], the authors estimated a secondary attack rate for the nosocomial spread of SARSCoV-2 between 0.73%-2.93%. They emphasised that the low attack rates characterised the beginning of the epidemics (the data in [19] focused on cases between January and April 2020), with higher attack rates expected if the epidemics would continue. They also emphasised that in Taiwan the nosocomial attack rates were very low, due to staff training and increased awareness as a result of previous experiences with SARS.

Based on these estimates, for our study we assume that each individual has a probability of 5% of becoming infected each day due to interactions with hospital staff and/or sharing the bathroom. In Figure 8 we see, for the first time in our simulations, that all patients have become infected in both 4-bed bays and 6-bed bays. Before considering interactions with staff members and sharing bathrooms, individuals at Beds 3 and 4 (4-bed bay) and individuals at Beds 4, 5 and 6 (6-bed bay) rarely became infected because the distance between beds was large enough to avoid the spread of the disease. However, if there are other ways of catching the disease, these patients will surely become infected. In fact, the probability of being recovered by *t* = 60 days is 1 for all the beds in both bays.

**Figure 8:**
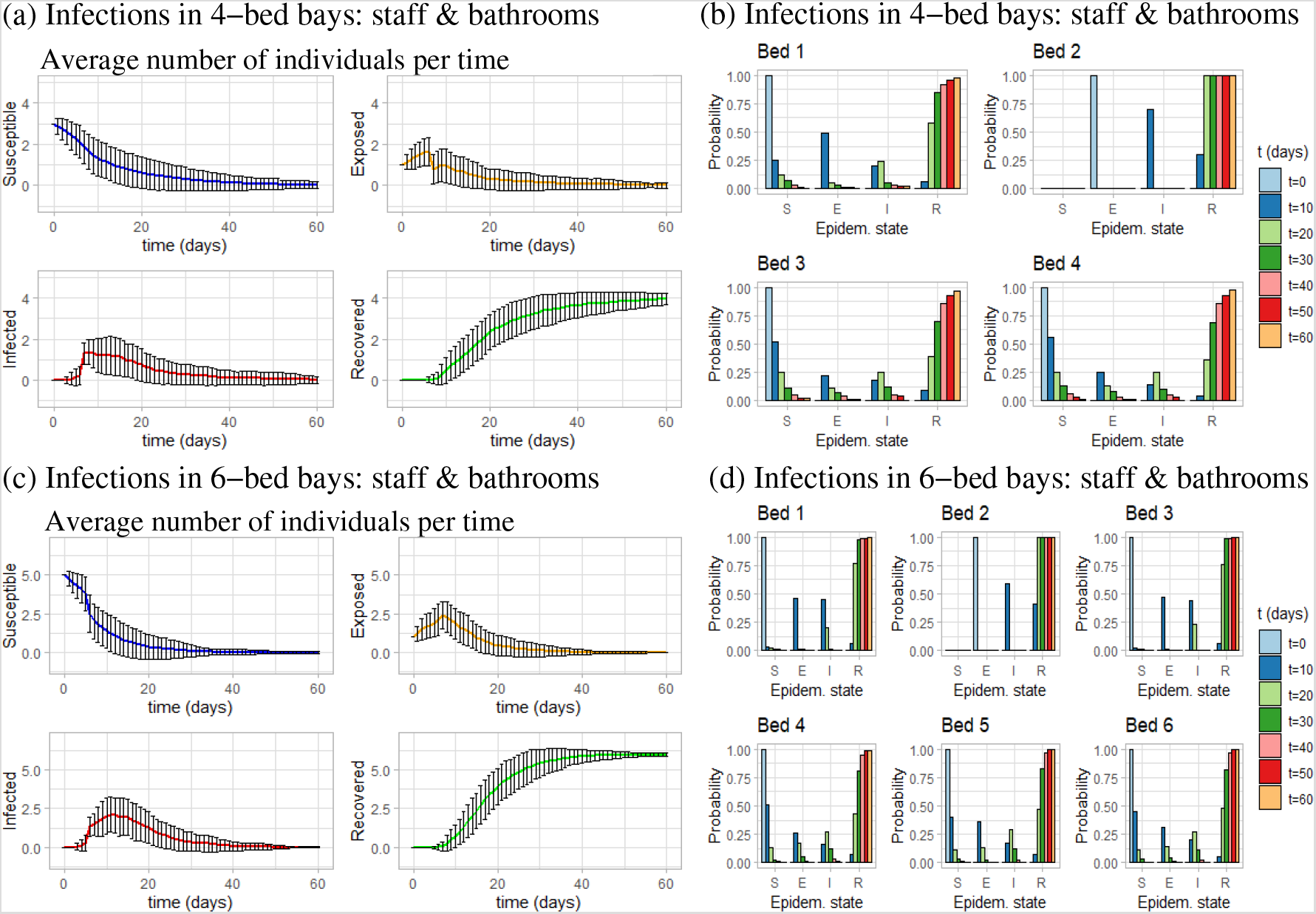
Infection spread throughout (a),(b) 4-bed bays, and (c),(d) 6-bed bays, when we assume that each individual has a 5% probability of becoming infected due to interactions with staff members or sharing bathrooms. (a) Average number of individuals per time in a 4-bed bay; (b) Probability of being at each epidemiological state per bed, in a 4-bed bay. (c) Average number of individuals per time in a 6-bed bay; (d) Probability of being at each epidemiological state per bed, in a 6-bed bay. As before, we assume that 1*/γ* = 7 days, 1*/σ* = 5.2 days, *T_max_* = 60 days, and Δ*t* = 0.1.

### 3.6. Disease dynamics in single rooms

In order to investigate the impact of disease spread following interactions with hospital staff, we now place only one individual in a single-bed room (which has an en-suite bathroom). Thus, the only way the patient can become infected is through interactions with the hospital staff. Since in this case we cannot talk about distance between beds, for the probability of infection *β* we assume that at the end of each day there is a fixed probability (5%) of becoming infected if the individual is susceptible.

As stated in [6, 30], the probability of infection is reduced by 85% when wearing masks. Here, we assume that the probability of infection is 5% per day when staff are wearing masks. Then we assume that the probability of becoming infected when staff are not wearing masks is 90%. In Figures 9(a) and (b) we show the probability of an individual (in a 1-bed room) of being at each epidemiological state when staff are wearing masks (sub-panel (a)) or not wearing masks (sub-panel (b)). It is clear that the use of masks increases the probability of the patient remaining in the susceptible state: from 0% after 10 days when the staff are not wearing masks, to 13% after 10 days when the staff are wearing masks.

**Figure 9:**
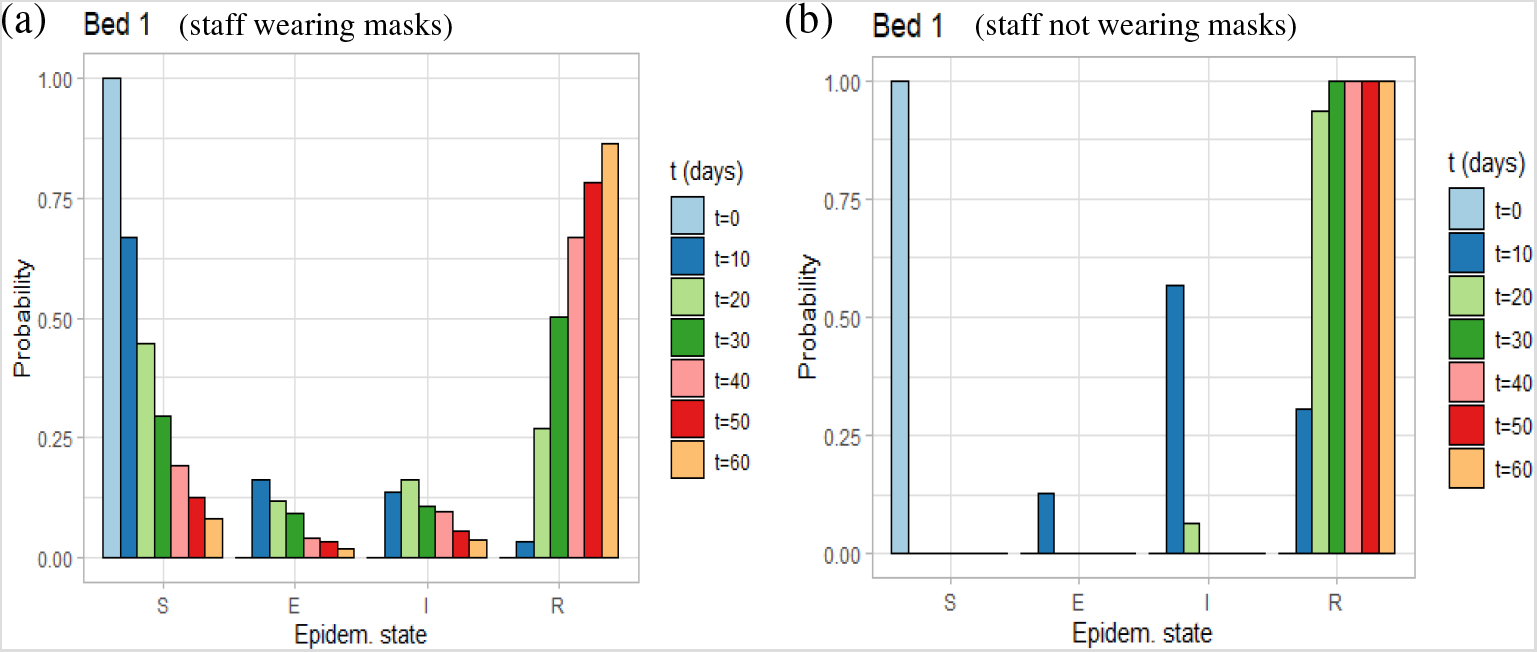
Single-bed hospital rooms: probability of being at each epidemiological state. (a) When hospital staff are wearing masks, infections may occur each day with a probability of 5%. (b) When hospital staff are not wearing masks, infections may occur each day with a probability of 90%. As before, the duration of incubation and recovery periods follows an Erlang distribution with means 1*/γ* = 7 days, 1*/σ* = 5.2 days.

## 4. The role of asymptomatic individuals

Since it has been shown that a large number of the people infected with Covid-19 do not present with symptoms [9], but they can spread the virus efficiently [27] next we investigate the effects these asymptomatic individuals have on model dynamics. To this end, we split the infected individuals into “infected asymptomatic” and “infected symptomatic”. Therefore, the epidemiological class *C_n,_*_1*,t*_ for each individuals can now take the following values: 0 (susceptible), 1 (exposed), 2 (infected asymptomatic), 3 (infected symptomatic) and 4 (recovered). Moreover, after the incubation period has passed, we randomly decide if the new individual is asymptomatic or symptomatic, with a maximum probability of 80% of being asymptomatic as suggested by various studies [20, 10, 9]. We assume that both exposed and infected asymptomatic individuals can transmit the disease, with the exposed individuals having a lower transmission probability (i.e., 20% lower, as considered also in Sub-section 3.4). We remove the infected symptomatic individuals the moment they become infected, and we remove the infected asymptomatic individuals after they test positive.

### 4.1. Results: the effect of frequent RT-PCR testing

Here, we compare the effects of running RT-PCR tests every day (Figure 10) or every 3 days (Figure 11) on hospitalised patients that have not been diagnosed with Covid-19. We see that it is better to run tests more often, as the probability of being susceptible (per bed) is higher when running tests every day (see panels (b),(d)). The results are similar for the 4-bay case and 6-bay case.

**Figure 10:**
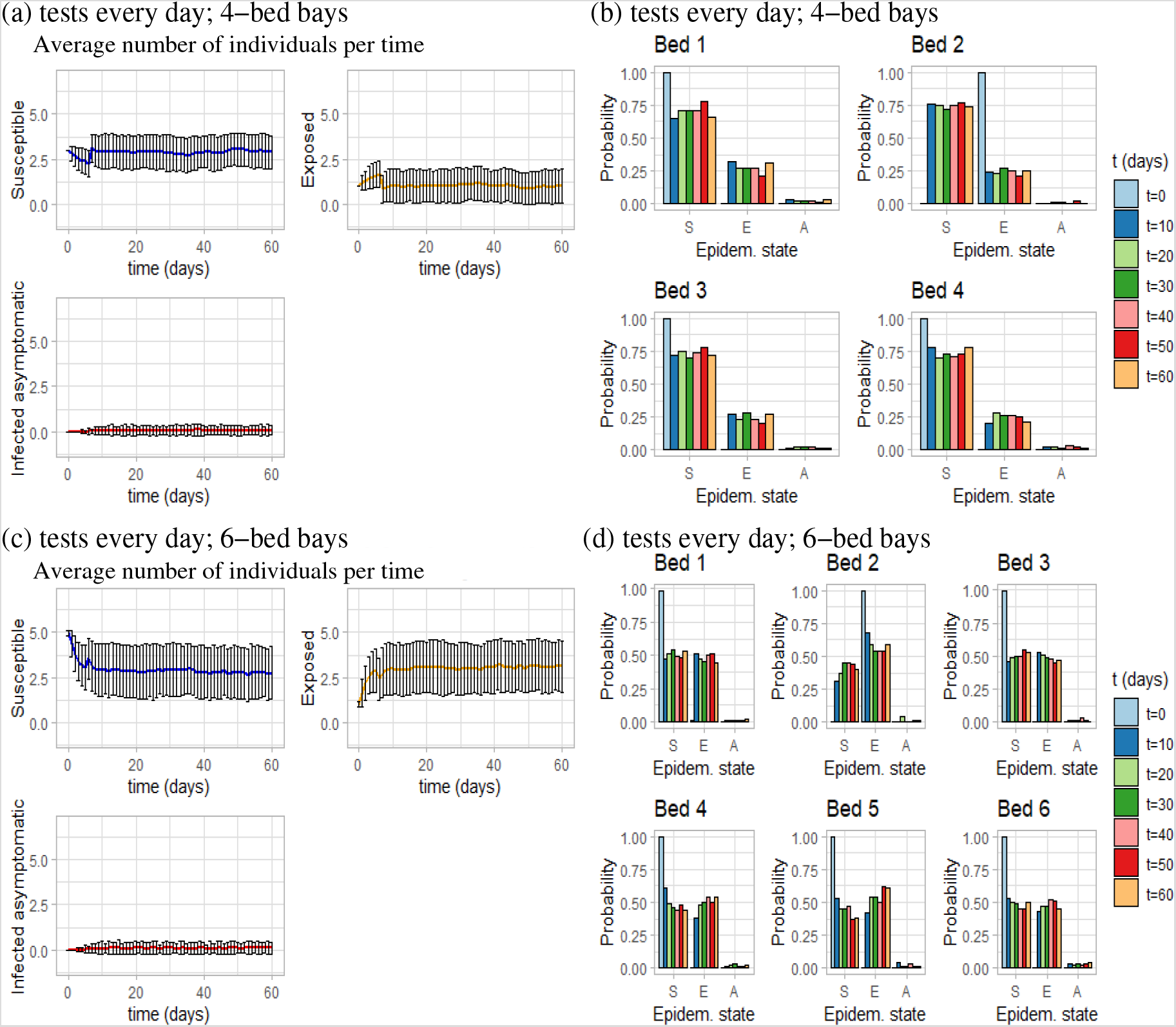
Disease spread when tests are run every day, and individuals have a 5% probability of becoming infected due to interaction with staff or by sharing bathrooms. (a) Average number of individuals per time, in a 4-bed bay. (b) Probability of being at each epidemiological state per bed, in a 4-bed bay. (c) Average number of individuals per time, in a 6-bed bay. (d) Probability of being at each epidemiological state per bed, in a 6-bed bay. As before, we assume that 1*/γ* = 7 days, 1*/σ* = 5.2 days, *T_max_*= 60 days, Δ*t* = 0.1.

**Figure 11:**
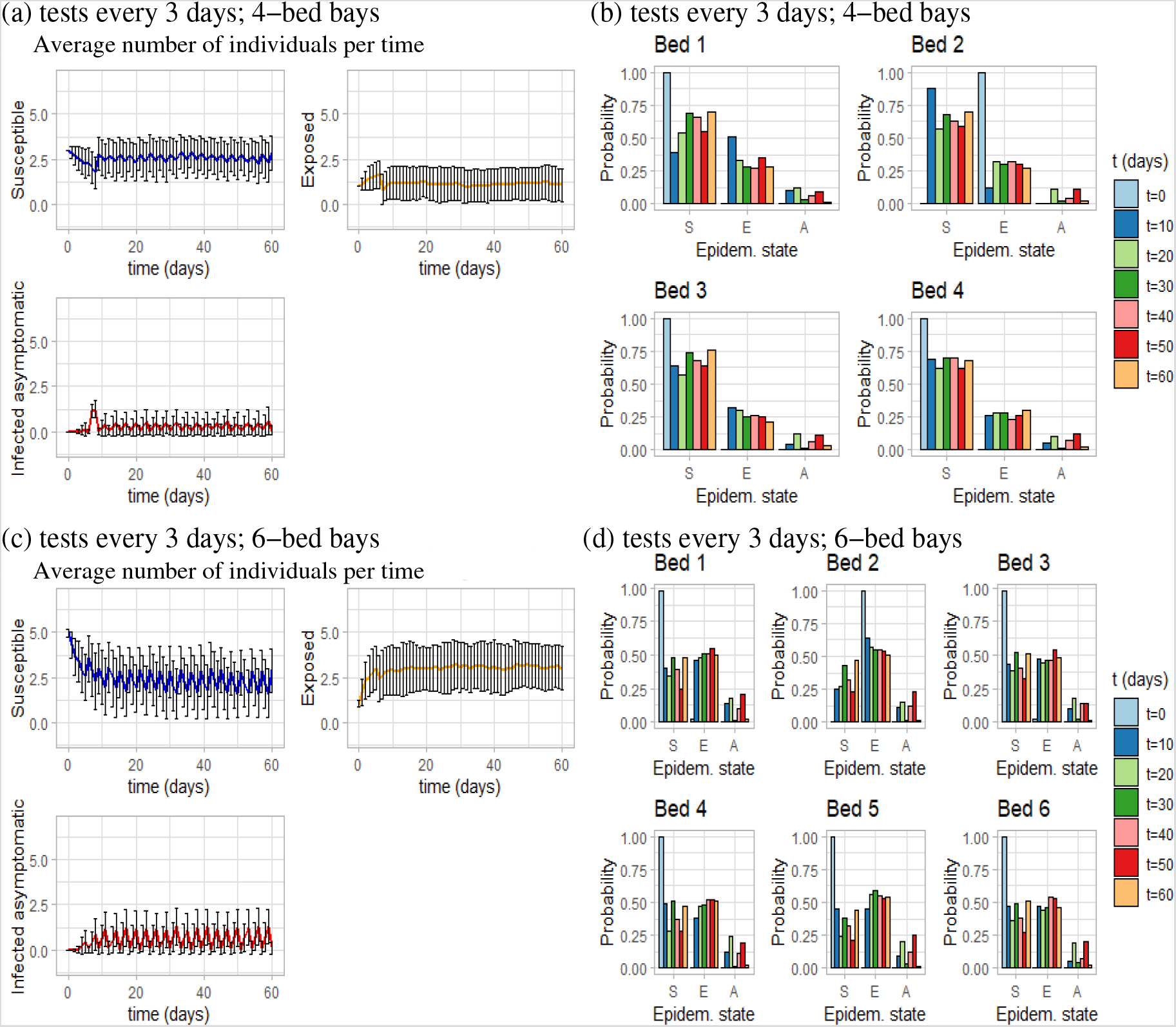
Disease spread when tests are run every 3 days, and individuals have a 5% probability of becoming infected due to interaction with staff or by sharing bathrooms. (a) Average number of individuals per time, in a 4-bed bay. (b) Probability of being at each epidemiological state per bed, in a 4-bed bay. (c) Average number of individuals per time, in a 6-bed bay. (d) Probability of being at each epidemiological state per bed, in a 6-bed bay. As before, we assume that 1*/γ* = 7 days, 1*/σ* = 5.2 days, *T_max_* = 60 days, Δ*t* = 0.1.

When running tests every 3 days (Figure 11), if we look at the infected asymptomatic individuals (in panels (a),(c)), sometimes we see two asymptomatic individuals in a bay. This is much higher compared to the case when tests are run every day, when the level of asymptomatics is zero (Figure 10(a), (c)). Moreover, for the case where tests are performed every 3 days, we note a periodic behaviour in the average number of individuals per time – caused by the removal of asymptomatics after they test positive.

## 5. Discussion

In this study we used modelling and computational approaches to investigate the question of the spread of an infectious disease through single rooms and shared hospital bays. This is an important aspect to be addressed due to the associated hospital costs of having single rooms or shared bays, and due to the very large numbers of patients that need to be hospitalised (with or without disease) in the context of a pandemic, as the one currently caused by SARS-CoV-2.

To investigate the probability that an infectious disease would spread among patients hospitalised in single rooms and in shared hospital bays with 4 beds or 6 beds, we used a simple network model that considered the probability that each bed had a susceptible, exposed, infected (symptomatic or asymptomatic) or recovered individual. With the help of this model we tested a variety of scenarios: the effect of placing the initial exposed individual in different beds inside the 6-bed bay (see Figure 4); the effect of varying the recovery and incubation periods (see Figures 5 and 6); the effect of removing infected individuals from the bays (see Figure 7); the effect of interaction with members of staff and sharing bathrooms, thus increasing the daily probability of infection (see Figures 8); the role of asymptomatic individuals and the effect of frequent testing (every day or every 3 days) that can be used to identify asymptomatic individuals (see Figures 10 and 11); and finally the effect of having only 1 patient per room, in which case the probability of infection depends only on interactions with staff members (see Figure 9).

We showed that patients hospitalised in single rooms have a higher probability to remain susceptible(i.e., not become infected) compared to patients in 4-bed bays; see Figures 9(a) and 8. We also showed that periodic testing combined with removing infected individuals from the bays reduces the probability of exposure and infection; see Figure 4.

## 6. Conclusion

We conclude our theoretical study by emphasising that healthcare associated infectious diseases seem to spread slower in single (1-bed) rooms versus shared bays. Moreover, the Covid-19 infection spreads slower in 4-bed bays compared to 6-bed bays. This predictive result is important since a 2015 report by National Institute for Health Research on the impact of single-room accommodation on patient and staff experiences [29] concluded that there was no clear evidence of a reduction in healthcare associated infections through single rooms: while many studies showed a reduction in infection rates, other studies showed no differences or higher infection rates. In our theoretical study we have shown that, from an epidemiological point of view, there are differences in the disease transmission when there is a single patient per room, or multiple patients per room. Moreover, when there are multiple room occupancies, it is important to keep the number of patients as low as possible (e.g., 4 instead of 6), to slow down the spread of infectious diseases. Regarding periodic testing, current guidance [44] advocates for routine testing of asymptomatic HCW every 7 days, depending on available testing capacity. Moreover, it is suggested that elective patients are to be tested for Covid-19 no more than 72 hours before admission, and cancer patients receiving long-course treatments should be tested every week [44]. Our simulation results investigated the effects of testing every 1–3 days. However, we emphasise that patient choice would need to be taken into account when developing policies for more frequent testing, as sampling has been reported to be uncomfortable. In addition, resources for testing would need to be increased in order for this to be feasible.

## Data Availability

All the data used for this work has been obtained from other published research papers.

## Acknowledgments (All sources of funding of the study must be disclosed)

It is a pleasure to acknowledge Mr. Wai-Lum Sung (graphic designer at the University of Aberdeen) for his assistance in the preparation of Figure 1.

The authors received no financial support for the research, authorship, and/or publication of this article.

